# Photovoice methodology to raise citizen awareness about the role of the gut microbiome in Non-Communicable Diseases: A pilot study

**DOI:** 10.1101/2022.06.17.22276351

**Authors:** Silvia Garcia, Sheyla Ordoñez, Enrique Carrillo de Santa Pau, Laura Judith Marcos-Zambrano

**Author notes:** Corresponding author: Laura Judith Marcos-Zambrano. These authors equally contributed to this work (Silvia Garcia, Sheyla Ordoñez).

## Abstract

**Background:** There is a lack of knowledge for the general public about gut microbiome implications on health and the ability of diet and lifestyle to change it, impacting the well-being and preventing the development of NCDs. Participatory actions with innovative digital methodologies such as photovoice motivate citizens to self-implement healthier habits and improve their health status.

**Methods:** We conducted a participatory action research initiative #PictureYourMicrobes (https://bit.ly/3wKuRYt), which consisted of a photovoice project accompanied by self-tracking citizen science activities (self-reported nutritional questionnaires and stool sample collection for microbial profiling). Learning outcomes related to recognizing the importance of nutrition over gut microbiome’s health and including prebiotics and probiotic foods in the diet were defined and evaluated through pre-and post-knowledge questionnaires.

**Results:** We recruited 70 volunteers to participate and set up six photovoice groups with homogeneous age and educational levels. Each group met online for four sessions composed of guided group discussions, where facilitators helped participants integrate their microbial profiling results with diet and nutritional habits. Participants took 156 photographs they thought best reflected how daily habits influence gut microbiota health. After contextualising and critically analysing the photographs and their reflections, they selected 64 photos sorted into four categories for a photobook (https://bit.ly/3LrUHEF) and exhibitions. They developed policy recommendations as a form of community-based solutions to improve gut microbiome health. Finally, we demonstrate that participants improved their knowledge about microbiota and its relationship with health through learning outcomes evaluation.

**Conclusions:** Participatory action research, such as photovoice, contributed to the awareness and adoption of new healthy habits, including the consumption of probiotics and prebiotic foods for a healthy gut microbiome. Moreover, it significantly impacted participants and provided a societal impact on other people through photobooks, pictures exhibitions, and digital media.

**Key questions:** *What is already known on this topic?:*

- Recent reports have shown that the gut microbiome (the complex of microorganisms that inhabit the gut) is altered in individuals with non-communicable diseases (NCDs).
- Citizens are unaware of the importance of the gut microbiome over health and the capability of diet and lifestyle modifications to modulate it and prevent the development of NCDs.

*What this study adds?:*

- The knowledge of the “own microbiome” allows people to reflect on their nutritional decisions and raise awareness about taking care of their microbiome and health.
- The use of participative action methodologies and co-creation activities in communication and dissemination allow participants to share their experiences and learnings with other citizens.
- Participative action methodologies used to teach complex health-related issues allows the transmission of knowledge from scientists to citizens.

*How might this study affect research, practice, or policy?:*

- Photovoice activity contributed to the awareness and adoption of new healthy habits, and participants became conscious of their nutritional habits through their reflections.
- Participatory action research activities allowed participants to propose solutions to maintain a healthy life and prevent NCDs critically and informally.
- Combining the results of these methodologies with social networks, digital photo books, and physical exhibitions amplifies the outcomes.
- Citizens developed policy recommendations as a form of community-based solutions to enhance gut microbiome health.

## Background

Non-communicable diseases (NCDs) are not transmissible directly from one person to another, including diabetes, cancer, cardiovascular, and chronic respiratory diseases [1]. Genetic factors and lifestyle habits like physical inactivity, unhealthy diet, and the harmful use of alcohol are tightly related to their development [1]. The World Health Organization manifests that 70% of deaths globally (∼41 million) are due to NCDs [2].

Recent reports have shown that the gut microbiome, the complex of microorganisms that inhabit the gut, is altered in individuals with NCDs [3]. Moreover, there is increasing evidence that obesity has a microbial component [4]. On the other hand, the gut microbiome has been shown to be responsible for personalised glycaemic responses of different people to the same meal [5]. It has a significant influence on postprandial lipemia [6].

Microbiome composition varies depending on diet, medication, exercise, and other environmental exposures [7]. In this regard, the diet has been signalled as a crucial factor in modulating microbiota, for instance, a low-fibre diet reduces the amount of beneficial microbiota and potentiates the growth of pathogenic bacteria. In contrast, a diet rich in fibre contributes to maintaining a “healthy gut microbiota” associated with increased diversity and functions, such as the production of short-chain fatty acids [8]. Hence strategies that might favourably modulate the gut microbiota to reduce the risk of many NCDs through diet and lifestyle changes are of great interest.

Despite the growing scientific research on the microbiome and its role in developing NCDs and the great body of knowledge supporting the relation between diet and host-microbiome interactions, there is limited consideration of the gut microbiome in dietary recommendations [9]. Moreover, there is still a lack of knowledge for the citizens about their implications on health and the ability of diet and lifestyle to change it, impacting the well-being and preventing the development of NCDs. Participatory Action Research (PAR) is a suitable strategy for approaching the population and understanding the lifestyle and health decisions from a citizens’ perspective to design more effective interventions that promote healthier environments [10–12].

Among PAR, photovoice has been gaining interest in social sciences [13–15], health care, and management of diseases [16,17]. It is defined as “a process by which people can identify, represent, and enhance their community through a specific photographic technique” [18]. On the other hand, applying “Citizen science” strategies, which includes a range of participatory models involving non-professionals as collaborators in scientific research, will benefit citizens’ empowerment and biomedical research [19].

Here we present “#PictureYourMicrobes”, a participatory action research project including Photovoice and self-tracking citizen science activities, with the primary objective of empowering citizens with risk factors for developing NCDs by raising awareness about the importance of taking care of the bacterial communities (microbiome) living in our bodies. This study reports on a participatory research initiative that brought together residents from different parts of Spain, nutritionists, civic centres, and academia to educate participants on microbiome health, raise awareness about health decisions and include the population in policy and decision-making.

## 2. Methods

This participatory action research project was conducted from April to September 2021, focusing on how lifestyle and nutritional decisions affect the microbiome’s composition and herein impact participants’ health.

### 2.1. Participants recruitment

We used a purposive sampling strategy to engage participants [20]. Our recruitment strategy was firmly based on social media, we use Facebook, Instagram, and Twitter channels from networks specialised in disseminating content related to nutrition and health. We also publicised the project on the platform: “Citizen Science Observatory” (https://ciencia-ciudadana.es/). Inclusion criteria were: 1) ages between 18-65 years, 2) live in Spain, 3) speak Spanish, 4) had no impediment to managing a digital camera, 5) not having a psychiatric disease or alimentary disorders, and 6) agreed to attend four group sessions for two months. Of the 200 people interested in participating, we selected 70 through stratified random sampling [20]. Study size was calculated with the pwr package Version 1.3 of Software R [21] assuming a change effect of 2 points as described in other works [22] with a standard deviation of 5, an error alpha of 0.05 and a power of 0.8, taking into account a dropout of 25% and 10% of poorly completed questionnaires.

### 2.2. Ethical Aspects and Data Processing

Protocols and methodology used in the present study comply with the ethical principles for research involving human subjects laid down in the Declaration of Helsinki (1964) and its modifications. The study was approved by the Research Ethics Committee of the IMDEA Food Foundation (PI-047; Approval date: March 11th, 2021). Participants were informed in detail about the different stages of the project both orally and in writing. The researchers collected signed informed consent prior to the first evaluation. This document included a specific consent to microbial profiling. Data compiled along the study was processed by applying dissociation criteria, making the volunteers’ data anonymous, in compliance with the current Spanish legislation (Organic Law 15/1999 of December 13th, on the protection of Personal Data).

### 2.3 Self-tracking citizen science tools

#### 2.3.1 Questionnaires and self-reporting data

We developed a 15-item knowledge questionnaire about the role of the microbiome in the development of non-communicable diseases and their relationship with healthy habits. Each participant’s knowledge score was computed by assigning 1 point to each correct response or 0 points, generating a 0−15. Higher scores represented a greater level of microbiome knowledge. The knowledge questionnaire was presented in session one before the project’s development and in session four after executing all the activities. A satisfaction questionnaire was also developed to evaluate to what extent participants engaged in the project. All items of the “satisfaction questionnaire” used a 5-point Likert response format (1 = strongly disagree to 5 = strongly agree for beliefs, and 1 = never to 5 = very often for attitudes). A validated food record of 72h consumption [23] in which participants had to sum up all food and drinks ingested during three days (2 weekdays and 1 Sunday or holiday) was delivered. Furthermore, the Global Questionnaire on Physical Activity of the World Health Organization was applied to assess the degree and intensity of physical activity [24].

#### 2.3.2 Microbial profiling

We collected stool samples for microbial profiling by 16S rRNA gene sequencing. Samples were collected in an OMNIgene•GUT collecting kit (DNA Genotek, Ontario, Canada) and sent by postal service in the next two days after sampling. DNA was extracted using the QIAamp DNA stool Minikit according to the manufacturer’s instructions (QIAGEN, Hilden, Germany). The variable V3 and V4 regions of the bacterial 16S ribosomal RNA gene (16S rRNA) were amplified from the faecal DNA and sequenced with the Illumina MiSeq platform (2 × 300).

#### 2.3.3 Photovoice procedure

We set up six Photovoice groups, homogeneous in age, educational level, and geographic region. Each group met online for four sessions, held every two weeks, and lasted approximately one and a half hours. Co-facilitators (an academic-based researcher and a nutritionist) facilitated the Photovoice groups, intervening only to explain the objectives of each session, involve everyone, and make sure that each participant was partaking. Each group session was recorded and transcribed. Sessions were divided into an initial group meeting, small group discussion sessions, and a final meeting.

In the first session, the research team introduced the project and the photovoice methodology. Participants performed an Initial knowledge questionnaire about the role of the microbiome in the development of non-communicable diseases and its relationship with healthy habits. Co-facilitators explained the procedure to collect the stool samples and provide explanations on how to fill the questionnaires and self-reporting data. In session 2, co-facilitators exposed the importance of diet and daily habits over gut microbe health, and participants were encouraged to take 2-3 photos that accurately represent the influence of their lifestyle on the composition of the microbiome. Also were provided with an adapted version of the SHOWED mnemonic method to guide discussions on why participants took that photograph and what it meant to them [18].The questionnaire included the following questions: 1) What does the photograph show?; 2) What is really going on here? 3) How does this relate to our lives and the health of the microbiome? 4) Why did you take this photograph? 5) What can we do about it? In the same session, participants received photography training. In session 3, participants discussed their photographs following SHOWED methodology and grouped all photographs into themes. In session 4, participants received their microbial profiling results and discussed the results considering the photographs and joining with their reflection after filling out the questionnaires and self-reporting data. Finally, participants developed policy recommendations to improve their diet and lifestyle habits to have better microbiome health and completed the knowledge questionnaire about the role of the microbiome in the development of non-communicable diseases and its relationship with healthy habits.

#### 2.3.4. Photobook and exhibition

An online photobook with Creative Commons Attribution Non-Commercial 4.0 International license was published in the ZENODO repository, including the selected photographs with their correspondent title and narrative given by the participants organised according to the proposed categories. An exhibition with photographs chosen was also prepared to be presented in different cultural and civic centres, allowing wider dissemination of the results, and increasing the study’s visibility.

### 2.5. Data analysis

Descriptive results are presented as the mean, interquartile range. Statistical analyses were performed using R software [25]. Qualitative and quantitative analysis of the diet was performed using the DIAL nutritional software v 3.15 (Alce Ingenieria, Madrid, Spain), and the calculated Healthy Eating index [26] was delivered to participants. Likert questionnaires were analysed using the Likert package in R software [27]. For the microbial profiling, data were processed using the Quantitative Insights Into Microbial Ecology program (QIIME2) [28] and annotated with the SILVA v.132 16S rRNA gene reference database [29]. The relative abundance of each ASV and alpha diversity (Shannon, Chao, and 211 Simpson indexes) were calculated using the phyloseq R package [30]. A final report was provided for all participants showing the main phyla, the *Firmicutes/Bacteroidetes* ratio, and the top 10 genera present in the sample. Statistical analysis was performed with the Kruskal-Wallis test or an independent Student’
ss t-test, being *P* < 0.05 values considered statistically significant.

### 2.6 Patient and public involvement

Participatory action research and citizen science are based on public participation. In our research, volunteers joined the focus group sessions, conducted the photovoice project, and collected data and stool samples. All the volunteers gave written informed consent and were informed about the burden of the intervention and the time required to participate. Participants were involved in the focus group discussions and contributed by creating recommendations and measures that were extracted to solve the existing problems in the diet and lifestyle of the population and promote positive behaviours and achieve a healthy microbiota. Moreover, they co-create a photobook and an itinerant exhibition allowing the dissemination of their experiences and learnings with other citizens, restarting the knowledge process. Finally, participants were informed about their results by a personalized report and were involved in several discussions related to the topic.

## 3. Results

Up to 200 people showed interest in participating in the project; we selected, through stratified random sampling, 70 volunteers that match the inclusion criteria. Finally, sixty-one volunteers completed all the stages of the project. A broader description of participants’ sociodemographic characteristics is presented in Table 1. Participants were aged 21–58 (mean age, 34), 85% females and 9% males, and most had higher education (85%). We have participants from sevens regions of Spain (Comunitat Valenciana, Cataluña, Galicia, Andalucía, Aragón, and Castilla la Mancha) however up to 79% were based in Madrid. Regarding the presence of risk factors for developing NCDs, we found that 34% of the participants were overweight/obese, 20% of people smoke, 20% have a poor diet according to the Healthy Eating Index [26], 23% have low physical activity according to the IPAQ questionnaire [24], and 11.4% has other risk factors for developing NCDs (hypercholesterolemia, hypertension, asthma, high sugar blood levels).

**Table 1.** Participants’ demographics and nutritional parameters were obtained after analysis of the food registry and IPAQ questionnaires.

| Sociodemographic characteristics |  | N=61<sup>a</sup> |
| --- | --- | --- |
| Age (years) |  | 34 (21,58) |
| Educational level<sup>b</sup> | Secondary education | 6(9.8%) |
|  | Tertiary education | 3(4.9%) |
|  | Bachelor's or equivalent | 19(31%) |
|  | Master o superior | 33(54%) |
| Residence | Madrid community | 48(79%) |
|  | Other communities | 13(21%) |
| Sex | Male | 9(15%) |
|  | Female | 52(85%) |
| BMI (kg/m<sup>2</sup>) | Normal range | 40(66%) |
|  | Overweight/Obese | 21(34%) |
| Smoking |  | 12(20%) |
| Physical activity <sup>c</sup> | Low | 14(23%) |
|  | Medium | 22(36%) |
|  | High | 25(41%) |
| Healthy Eating Index <sup>d</sup> | Poor | 11(20%) |
|  | Fair | 12(21%) |
|  | Good | 18(32%) |
|  | Very Good | 11(20%) |
|  | Excellent | 4(7.1%) |
|  | NA | 5 |

<sup>a</sup>median (range); n(%). <sup>b</sup> According to the International Standard Classification of Education.
<sup>c</sup> according to the IPAQ questionnaire [24], <sup>d</sup> According to Healthy Eating Index [26].

### 3.1 Self-tracking citizen science tools

In order to obtain a profile regarding the diet, physical activity and sociodemographic characteristics of each participant, we provide self-reporting questionnaires to record data. We encourage participants to use the responses of the food registries as a tool to enrich their photovoice narrative and intensely elaborate a reflection on their habits. On the other hand, we perform microbial profiling of the participants’ gut microbiota (n=61) trough sequencing the 16s rRNA gene, we calculate the relative abundance of the phylum present in each sample, the *Bacteroides/Firmicutes* ratio and the most common genera present. We prepare an individualized report for each participant and discussed the microbial results in session 4. All the personal questions regarding the analysis were solved to increase self-awareness about the gut microbiota and its relation to diet and lifestyle habits.

We asked participants to include their acquired knowledge in photovoice narratives. Participants’ reflections suggest that microbial profiling and self-reporting questionnaires effectively help realize what habits need to change or maintain. e.g.:*“Thanks to my participation in this project, I have been able to know myself better in terms of my lifestyle. Through all the activities carried out and particularly by the microbial profiling… It has made me reflect that I should change habits”*.

### 3.2 Photovoice project

Participants took 156 photographs during the entire Photovoice project and selected 64 as the ones best reflecting their food and lifestyle environment impacting gut microbiome health. After discussion and participative communication, they grouped photographs into four themes: (1) balance, (2) foodie, (3) mindful eating, and (4) wellness. Participants’ results are presented according to these themes, following participants’ selected photographs, their SHOWED-based narratives, and other group members’ related discussions.

#### 3.2.1. Theme 1: Balance

One of the most relevant topics that arose during discussions in the six groups was the importance of mental health and emotional well-being to achieve vibrant health. According to the World Health Organization (WHO), “health” is a state of complete physical, mental and social well-being, not merely the absence of disease [31]. Participation in leisure activities, having a pet, and potentiating good healthy habits were pointed out as ways to have a balanced life that, according to our participants, will be directed to better gut microbiome health.

Participants who worked emphasised that schedules and workloads often do not allow them to have good nutritional habits, and they were seeking balance. For example, a participant Supplementary Fig 1a: *“A sedentary job has no long-term benefits to health”… “I feel additional stress, which has made my stomach not work the same, my body and my mood are resentful*.*”* Also, participants emphasised the importance of leisure time to have better mental health and, herein, better gut health. *“Having time to dedicate it to yourself is very important. It helps you to have a better mood and decrease stress and anxiety levels*.*”… “It is an important factor for good health, both mental and physical*.*”*. In addition, having pets was suggested as an important factor for boosting well-being by helping, on the one hand, mental health and, on the other hand, influencing gut-microbiota, as an example Supplementary Fig 1b: *“Pets bring happiness and company. In particular, regarding the microbiome’s health, it is proven that having pets, especially in the first years of life, when the microbiota is developing, favours diversity and protects against diseases related to immunity, such as allergies or asthma*.*”*

#### 3.2.2. Theme 2: Foodie

Spanish cuisine has great importance in the way of communicating in Spanish society, which plays a relevant part in celebration and family environments. These events, generally, happen in a relaxed atmosphere with family and friends where healthy food is not prioritised. Participants gave relevance to moderate eating and finding the “healthier option” when celebrating and having leisure time with family and friends.

Participants showed that sometimes eating outside is not the healthier option. Supplementary Fig 2a: *“We tend to eat portions with amounts of fat, salt, flours*… *much larger than we would eat at home*.*”* … *“I took this picture to show the other side of the coin when I have no time, or I don’t want to spend hours and hours preparing food*.*”*. On the other hand, some participants pointed out the idea of preparing the same dishes found in a restaurant but with healthier options and homemade. e.g.: Supplementary Fig 2b: *“A homemade dish from Chinese restaurants in Spain*.*”. “Rice is a very nutritious food containing resistant starch. Resistant starch is an excellent food for the microbiota as it becomes prebiotic. It has a low glycemic index which reduces the inflammatory effects of the intestine”… “When I became more interested in leading a healthy lifestyle, I missed fast food dishes. Then I decided to make these meals at home in their healthy version*.*”*

#### 3.2.3. Theme 3: Mindful eating

Diet and nutrition were among the most critical factors pointed out by participants. They proposed the theme of Mindful eating as the idea of paying attention to an eating experience and witnessing the emotional and physical responses that take place before, during, and after the eating experience. Furthermore, to conscientiously take care of the gut microbiome by including fibre probiotics and a balanced diet.

In Supplementary figure 3a, we showed a bowl of kefir. The participant who took this picture commented: *“Our microbiota is made up of lots of living bacteria that take care of our intestinal health and prevent the development of diseases. When we take food with natural probiotics, we directly introduce those living microorganisms that are very beneficial for our health and directly affect our microbiota*.*”*. On the other hand, the acquisition of fresh fruit and vegetables as a source of fibre was also relevant. Supplementary Figure 3b: *“The relationship between microbiota and food is very close, and local food can help us achieve a healthier, more sustainable diet to improve it*.*”… “Local foods have great virtues; the nutritional power of freshly cut fruit and vegetables, its texture, and the fact that it maintains all its properties and is ecological… unlike the food that we can find in any supermarket and/or shopping centre*.*”*

#### 3.2.3. Theme 4: Wellness

Participants specified that exercise was essential for having better gut health. For many people, exercise is their way of disconnecting from their daily routine, work, and stress. Participants pointed out that physical activity *e*.*g*.: going to the gym, outdoors activities, or doing indoor exercise, is closely related to gut microbiota and well-being.

In Supplementary figure 4a, a participant said: *“Sport is essential to have a healthy life. Whether or not it is directly related to the microbiome, it has been shown to have a very beneficial impact; it reduces stress, improves sleep, increases our metabolism and muscle mass, densifies bone mass, and can be linked to a good relationship with food*.*”…” Physical exercise not only makes us feel good but also influences the way we metabolize macronutrients healthily*.*”*. Other participants also showed the importance of using alternative ways to commute to work, such as walking and using the bike, reflected in the Global Physical Activity Questionnaire, as a way to do exercise, *i*.*e*., *“How we move around the city is a choice that affects both our health and the environment*.*”. “A moderate exercise outdoors, such as regularly commuting by bike, makes us more active, positive and improves our physical condition and metabolism, including our microbiota*.*”*. Overall there was a consensus on the importance of creating a routine to exercise and improve health: Supplementary Fig 4b: *“Regular physical activity can increase the growth of beneficial bacteria for our body”… “I try to exercise every day, doing yoga, doing static rowing or walking in the field*.*”*

### 3.3 Knowledge obtained and satisfaction

Learning was evaluated through pre-and post-questionnaire questions. The percentage of correct answers after and before the project’s participation is recorded in Table 2. We define four learning outcomes: (i) State the concept of the microbiome. (ii) Identify differences between prebiotics and probiotic foods. (iii) Recognize the importance of nutrition over the microbiome and health. (iv) Discuss the implication of daily nutritional decisions over gut health. After project execution, participants could recall the microbiome concept, as seen in the increased percentage of correct answers in question 1 (Table 2). Moreover, they summarised different aspects related to the microbiome, as seen in questions 7, 8 and 9 (Table 2). Some of the reflections after the focus group activities also showed the accomplishment of the learning outcome. For example, a participant said: “*I have learnt that we have many types of bacteria in the intestine, that each one regulates different things*… *but there are no more important bacteria than others*… *the most important thing is to have a balanced amount of each one of them*”. Also, participants comprehend the difference between probiotics and prebiotics, as seen in the percentage of correct questions numbers 2, 3, 11 and 12 (Table 2). As seen in the focus group discussion reflections, participants understood and discussed the importance of daily nutritional decisions over gut microbiome health. A participant states: *“The gut microbiome comprises a very diverse group of microorganisms with different metabolic functions, which are directly linked to our behaviour and nutritional habits. We are what we ea*t”. Other participants said: “*Diet and lifestyle have a great influence on the good condition of our microbiota*… *so it is never too late to make changes for improvement*”, *“I have learnt that a balanced microbiota is essential for physical and mental well-being. A diet that takes care of our microbiota is life insurance*”. Finally, we achieved an overall increase of 0.72 points in the mean results of the questionnaires (*P* < 0.001).

**Table 2.** Recommendations and interventions developed by participants after photovoice project execution. All recommendations are shown cording to the photovoice theme.

| Recommendation | Interventions | Theme |
| --- | --- | --- |
| **Increase the quality of leisure time moments**<br>To reduce stress caused by the high pace of daily life, work, and lack of rest. | Socializing to maintain an optimal mood and keep the brain active.<br>Getting 8 hours of sleep.<br>Going for a walk in a pleasant environment. | *Balance* |
| **Don't give up the food we like**<br>To maintain good social health by avoiding daily monotony. | Eating out with family and friends.<br>Consuming "unhealthy" foods occasionally.<br>Cooking with healthier ingredients and at home. | *Foodie* |
| **Implement nutritional education programs**<br>So that the new generations become aware of the importance of a healthy diet in their microbiota. | Providing nutrition education classes in schools.<br>Teaching the smallest healthy eating habits in the family environment. | *Mindful Eating* |
| **Increased availability of fresh fruits and vegetables**<br>To achieve a more varied and healthy diet by taking advantage of the seasonality of food. | Choosing zero-kilometer products in local internet stores.<br>Creating organic gardens at home.<br>Replacing prepared or ultra-processed foods with fresh products. | *Mindful Eating* |
| **Increase consumption of prebiotics and probiotics**<br>To improve and strengthen the health of our microbiota. | Making at home Kefir, Kimchi, or Kombucha.<br>Attending prebiotic food courses. | *Mindful Eating* |
| **Encouraging non-motorized means of transport**<br>To increase daily physical activity. | Promoting movement on foot.<br>Making use of the cycling in urban areas. | *Wellness* |
| **Promote sport**<br>For the sedentary, motivating citizens to practice collective or individual exercise. | Performing different exercises at home with the necessary equipment.<br>Attending classes or scheduled activities. | *Wellness* |

To evaluate to what extent participants have enjoyed partaking in the project, we develop a questionnaire with a Likert scale regarding changes in habits and recommendations after the project’s participation and global satisfaction after participation in the project (Figure 1). We found that up to 95% of the participants were satisfied with their involvement in the project, and 93% would recommend participating in it. Up to 95% increased their interest in healthy eating, and 95% of the participants stated that they became more aware of the importance of maintaining a good diet and having healthy lifestyle habits to prevent diseases. Finally, 86% found all their doubts about the nutrition-microbiome relationship solved, and 79% found the photovoice project strongly useful.

**Figure 1.**
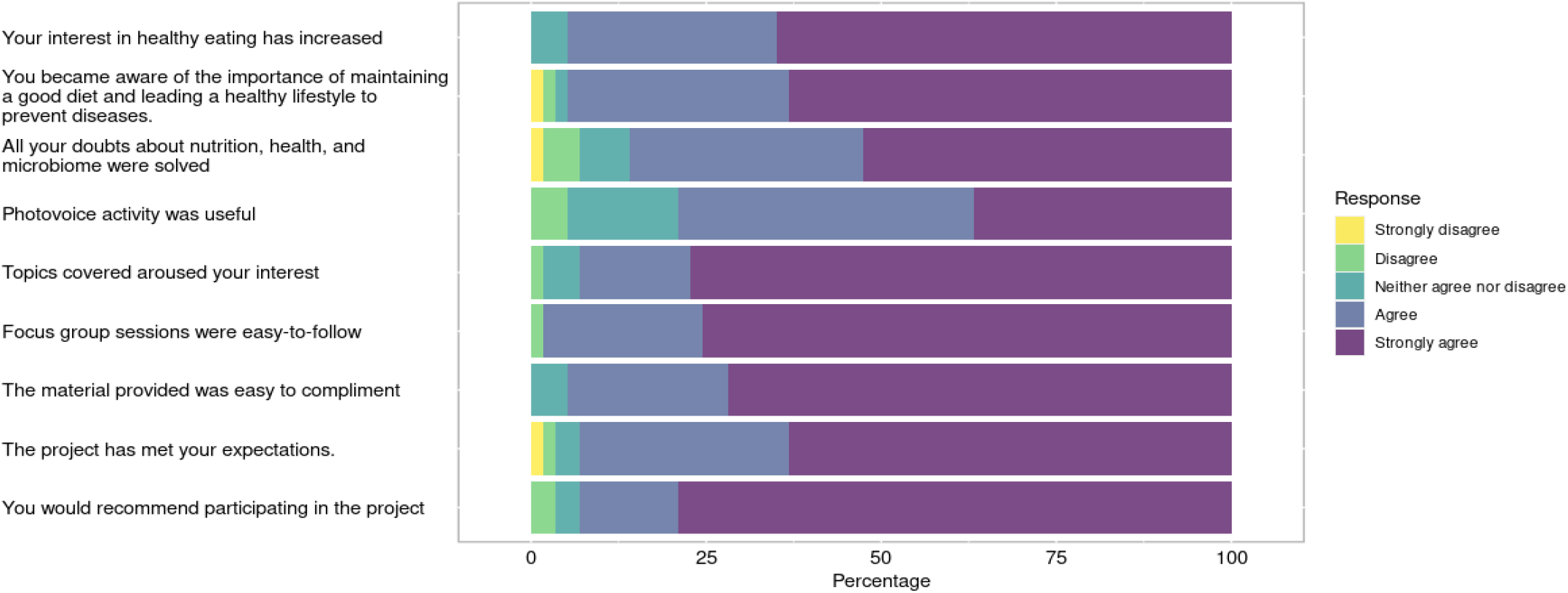
Satisfaction questionnaire regarding project participation, changes in habits, and recommendations. Answers were presented on a ert scale from 1 (strongly disagree) to 5 (strongly agree).

### 3.4 Recommendations and interventions

Following the logical framework methodology of the project, after the intervention in the sessions, we carried out an analysis and categorisation of the photographs based on the problems of the environment and positive aspects of the daily life of the participants. Table 3 shows the recommendations and measures extracted to solve these existing problems in the diet and lifestyle of the population and promote positive behaviours and achieve a healthy microbiota. In the category “Balance”, participants suggest increasing the quality of leisure activities to disconnect from daily stress by socialising with friends and family and recalling the importance of good sleep. In the “Foodie” category, participants state the importance of having good nutritional decisions when going out with family and friends. In the “Mindful eating” category, participants advise to increase the consumption of probiotics and prebiotic foods and prompt the elaboration of homemade probiotic food such as kefir and kombucha. On the other hand, they recognise the importance of eating fresh vegetables and fruits and encouraging organic food consumption, creating urban gardens, and consuming kilometre zero products. Finally, for the category “Wellness”, participants promote sports and motivate people to use non-motorized transport.

**Table 3.**
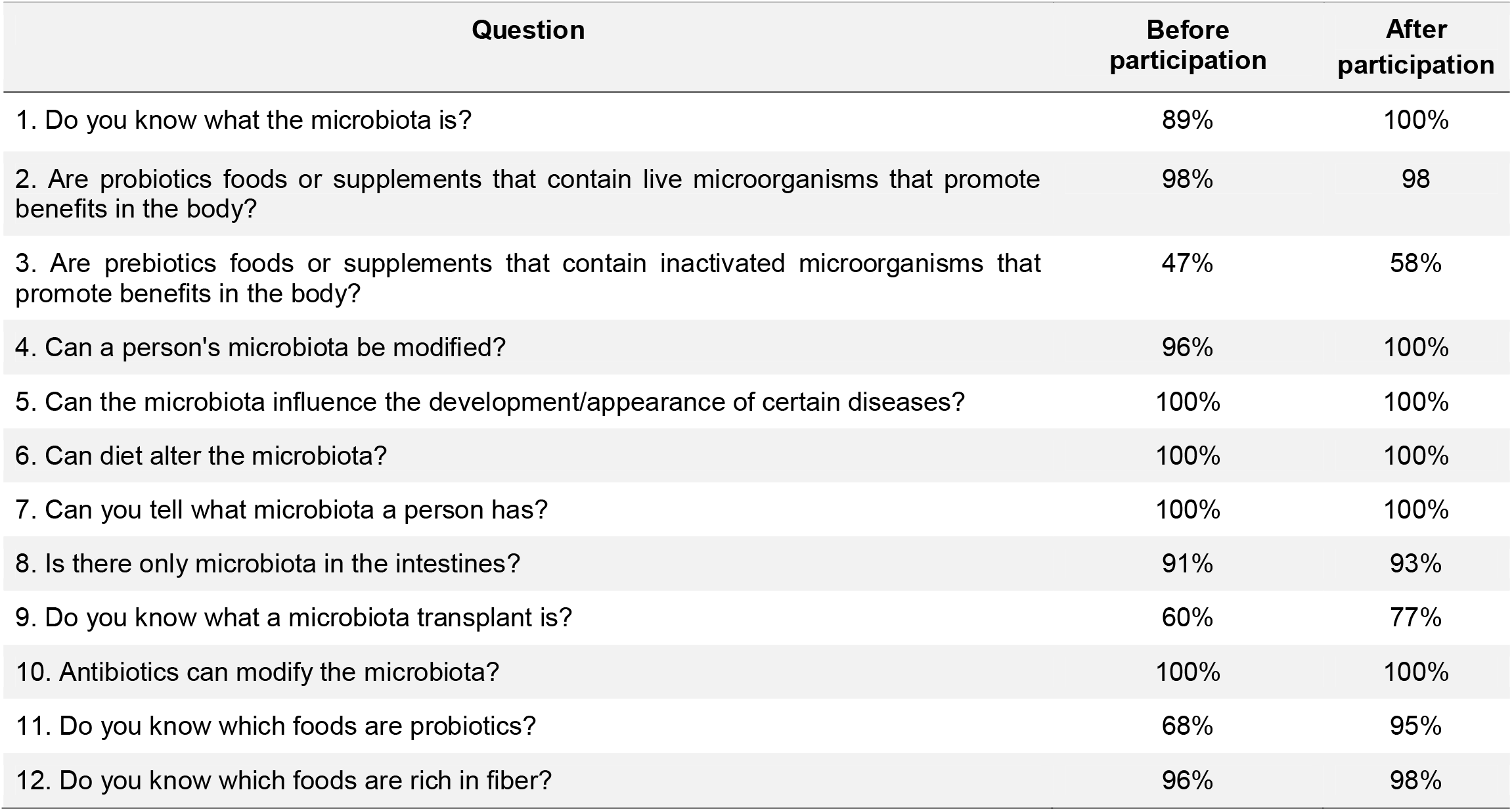
Percentage of correct answers after and before participation in the project.

| Question | Before participation | After participation |
| --- | --- | --- |
| 1. Do you know what the microbiota is? | 89% | 100% |
| 2. Are probiotics foods or supplements that contain live microorganisms that promote benefits in the body? | 98% | 98 |
| 3. Are prebiotics foods or supplements that contain inactivated microorganisms that promote benefits in the body? | 47% | 58% |
| 4. Can a person's microbiota be modified? | 96% | 100% |
| 5. Can the microbiota influence the development/appearance of certain diseases? | 100% | 100% |
| 6. Can diet alter the microbiota? | 100% | 100% |
| 7. Can you tell what microbiota a person has? | 100% | 100% |
| 8. Is there only microbiota in the intestines? | 91% | 93% |
| 9. Do you know what a microbiota transplant is? | 60% | 77% |
| 10. Antibiotics can modify the microbiota? | 100% | 100% |
| 11. Do you know which foods are probiotics? | 68% | 95% |
| 12. Do you know which foods are rich in fiber? | 96% | 98% |

### 3.5 Photobook and Exhibitions

We developed activities to favour citizen engagement and help disseminate the presented results. First, we designed a photobook including 64 photographs selected by the participants with their title and correspondent narrative. The Photobook was published online in the zendo repository under a Creative Commons Attribution Non-Commercial 4.0 International license on September 2021 and is free downloadable (https://zenodo.org/record/5682942/files/FOTOLIBRO_licencia.pdf?download=1). Then we organise an itinerant exhibition to be held at civic centres to disseminate the results obtained and was visited with more than 1,400 people.

## 4. Discussion

The integrated use of photovoice and citizen science activities is a novel approach to comprehending social and environmental factors that influence gut microbiome health. Photovoice provided information on how participants perceived their environment. Whereas citizen science activities allow participants to increase their understanding of their dietary choices, quantify physical activity performance, and microbial profiling at a glance. These data allowed participants to identify strengths and weaknesses regarding their habits and construct healthier opportunities considering the gut microbiome health.

A significant body of literature supports the use of photovoice for social sciences [13–15,32] and the management of diseases such as diabetes [16,17]. On the other hand, citizen science approaches in biomedical sciences have demonstrated great benefits, particularly in “data collection” projects in which people with access to their data increase interpretive fluency [19,33]. The research advances between nutrition and microbiome disciplines validate current dietary guidelines. However, there is a need to incorporate knowledge on the molecular basis by which nutrients impact host-microbiome interactions to improve human nutrition potentially [9]. PAR activities used to teach these interactions may help transmit knowledge to the citizens and influence their awareness about adopting healthy habits to boost health. As far as our knowledge, this is the first study in which photovoice and citizen science are combined to increase the understanding of gut microbiome health and improve lifestyle habits.

Due to our recruitment strategy, the cohort of participants was homogeneous, comprised mainly of young women with high educational levels with great interest in health, weight loss and learning about the microbiome, and users of social media channels. We focused on determining in our cohort of participants risk factors associated to the development of non-communicable diseases according to WHO [2], namely: living with overweight or obesity, unhealthy diets determined with a score of <50% in the HEI, smoking habits, low physical activity as determined by the IPAQ questionnaire, and other factors such a diagnosed hypertension or hypercholesterolemia. We found up to 67% of the participants with risk factors for developing NCDs, particularly 36% of the participants has only one risk factor associated to NCDs, and 31.1% has two or more combined factors to develop NCDs. Overall, these characteristics directed the participants’ interest to some points that were the main themes of the photovoice project, recalling stress, pet-owning, diet, and exercise as critical issues affecting their gut microbiota and, to a greater extent, their global health.

Stress is a nonspecific response of the body to any demand imposed upon it [34]. It can hardly be avoided in the present competitive world. Stress has been associated with harmful effects on physical health and has negative implications for the endocrine, immune and central nervous systems [35]. It can also affect the state of the intestinal barrier [36], has been associated with an increase in gut permeability [37] and impact the gut microbiota-brain axis [38]. Participants recognise that the correct handling of stress is necessary for better global health, such as doing outdoor activities, practising sports, enjoying with family and friends, and having pets.

Besides the beneficial effect on mental health and well-being of pet ownership [39,40]. It has been demonstrated an association between having pets and the microbiome [41]. Exposure to pets is known to decrease the rate of atopic and allergic disease [42,43] and acts as much on the adult gut as altering the maturing gut microbiome during infancy [44,45].

Finally, participants recall the importance of exercise and sports over gut microbiota health. It has been demonstrated that exercise training improves gut microbiota profiles and reduces endotoxemia [46]. Moreover, differences in the gut microbiome can be observed among professional athletes and less active subjects [47], suggesting a clear relationship between exercise and gut microbiome composition.

Our intervention showed an impact on participants by increasing their knowledge of the topic by 0.72 points, moreover up to 95% of the participants stated that they became more aware of the importance of maintaining a good diet and having healthy lifestyle habits to prevent diseases, as shown in our questionnaires. Finally, we demonstrate that the application of participatory action research is a way to include the population in policy and decision-making and empower participants by improving knowledge and making better health decisions.

Despite the extraordinary interest aroused by the study, we had a dropout of nine participants. The photovoice project required active participation and significant commitment from the participants. Although we developed the photovoice sessions online, sessions were time-consuming besides self-reporting questionnaires, and narrative construction and reflection for the photographs demanded some time that some participants did not have. Moreover, two participants had to leave the project due to technical issues. Some studies suggest that a dropout of >20% of the participants will cause severe threats to validity [48]. In our case, more than 87% of the participants finished the project, and 93% will recommend participating in it, demonstrating a high commitment level and satisfaction.

Finally, we showed that our approach contributed to the awareness and adoption of new healthy habits, including the consumption of probiotics and prebiotic foods for a healthy gut microbiome. Moreover, it significantly impacted participants and provided a societal impact on other people through photobook, pictures exhibitions, and digital media.

## Data Availability

All data generated or analysed during this study are included in this published article. Photobook is published under Creative Commons Attribution-NonCommercial international license (CC-BY NC) online in Zenodo repository.

https://doi.org/10.5281/zenodo.5682942

## Ethics approval and consent to participate

The study was approved by the Research Ethics Committee of the IMDEA Food Foundation (PI-047; Approval date: March 11th, 2021). Participants were informed in detail about the different stages of the project both, orally and in writing. Signed informed consent was collected by the researchers prior to the first evaluation.

## Consent for publication

Not applicable

## Availability of data and materials

All data generated or analysed during this study are included in this published article. Photobook is published under Creative Commons Attribution-NonCommercial international license (CC-BY NC) online in Zenodo repository with the DOI: https://doi.org/10.5281/zenodo.5682942.

## Authors’ contributions

Conceptualization, L.J.M.-Z. and E.C.d.S.P.; investigation, data curation, methodology, and formal analysis S.G, S.O and L.J.M.-Z.; writing—original draft preparation, L.J.M.-Z; writing—critical review and editing, E.C.d.S.P; funding acquisition, L.J.M.-Z. and E.C.d.S.P. All authors have read and agreed to the published version of the manuscript.

## Acknowledgments

We want to thank our 70 participants and those around 152 more people who signed up for our recruitment. To all who have followed and supported us on social media and continue to do so. To Paula Guiu for collaboration during all the stages of this project and Gloria Fuentes for the infographics designs for the photobook and exhibitions. To the following civic centers, in Zaragoza, for being the first to host our exhibition: Río Ebro, Teodoro Sánchez Punter (San José), La Almozara.

