## Supplementary material for "Photovoice methodology to raise citizen awareness about the role of the gut microbiome in Non-Communicable Diseases: A pilot study": Suppl. material

**Supplementary figure 1. (a)** Photograph reflecting stress and high workload: “Working from home, many hours of sedentary lifestyle and loneliness” Theme: Balance. **(b)** Photograph reflecting the importance of having pets to wellness: “The implication of having a pet with my microbiota”. Theme: Balance.

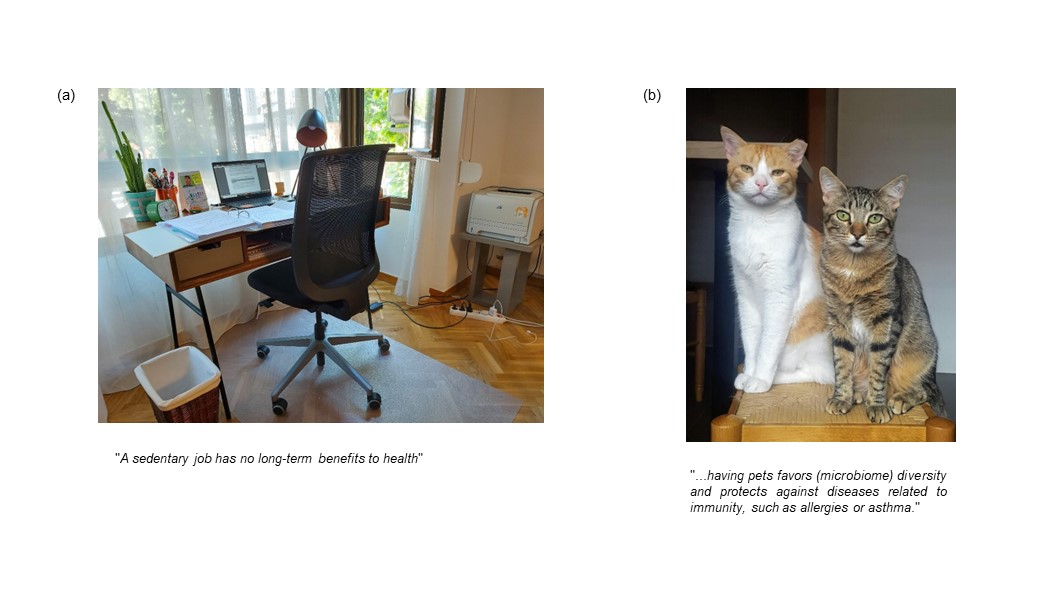

**Supplementary figure 2. (a)** Photograph reflecting unhealthy food elections: “Fast food, to calm hunger quickly and easily” Theme: Foodie. **(b)** Photograph reflecting the importance of better health decisions in leisure time: “Bowl with Asian style rice”. Theme: Foodie.

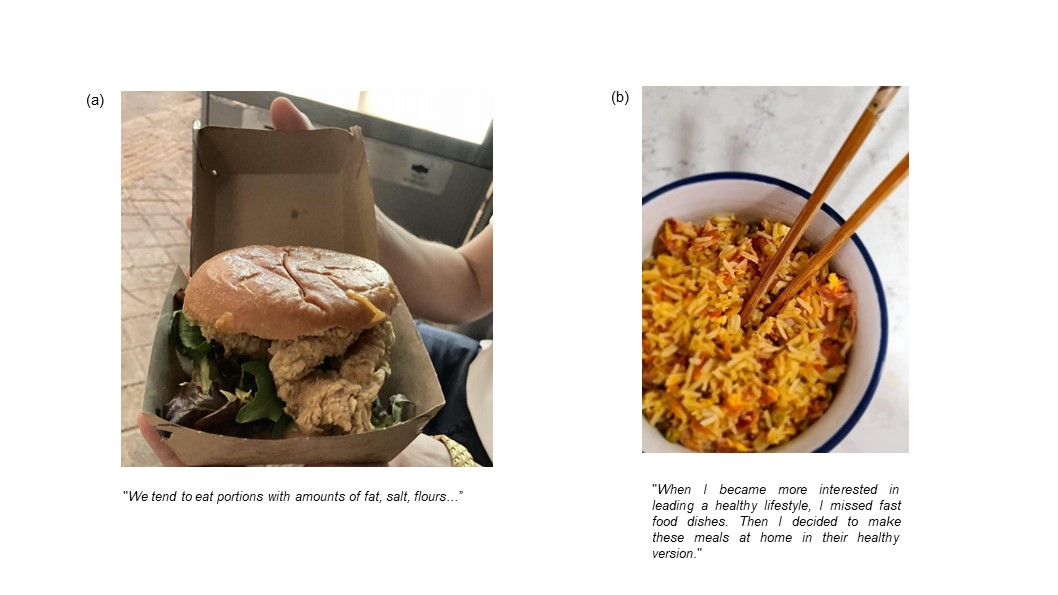

**Supplementary figure 3. (a)** Photograph of homemade probiotic food: “The superfoods are the ideal choice” Theme: Mindful eating (b) Photograph showing fruits and vegetables: “Local farmers who produce high-quality food”. Theme Mindful eating.

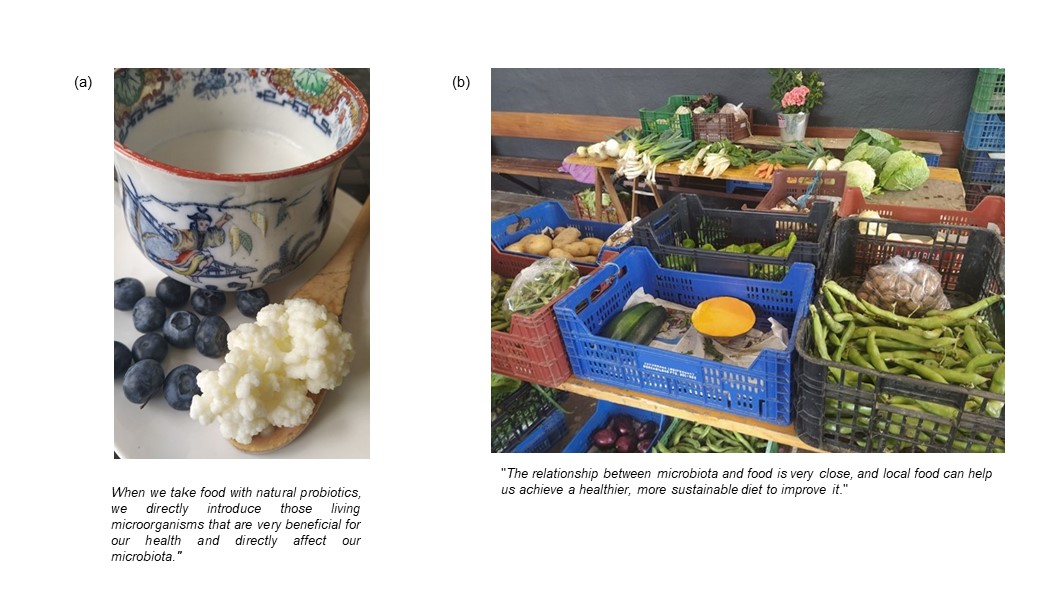

**Supplementary figure 4. (a)** Photograph reflecting an improvised homemade gym: “Gym at home, in an easy, practical and comfortable way” Theme: Wellness. (b) Photograph reflecting the benefits of doing exercise: “Yoga posture in a pleasant environment”. Theme: Wellness.

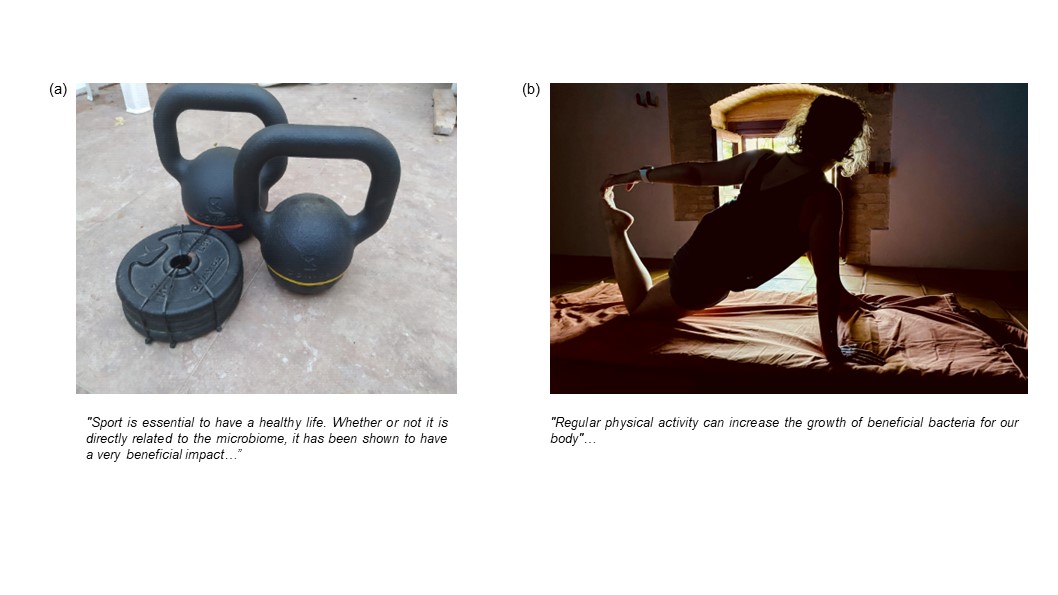
